# Post-stroke changes in brain structure and function can both influence acute upper limb function and subsequent recovery

**DOI:** 10.1101/2024.11.02.24316641

**Authors:** Catharina Zich, Nick S Ward, Nina Forss, Sven Bestmann, Andrew J Quinn, Eeva Karhunen, Kristina Laaksonen

## Abstract

Improving outcomes after stroke depends on understanding both the causes of initial function/impairment and the mechanisms of recovery. Recovery in patients with initially low function/high impairment is variable, suggesting the factors relating to initial function/impairment are different to the factors important for subsequent recovery. Here we aimed to determine the contribution of altered brain structure and function to initial severity and subsequent recovery of the upper limb post-stroke.

The Nine-Hole Peg Test was recorded in week 1 and one-month post-stroke and used to divide 36 stroke patients (18 females, age: M = 66.56 years) into those with high/low initial function and high/low subsequent recovery. We determined differences in week 1 brain structure (Magnetic Resonance Imaging) and function (Magnetoencephalography, tactile stimulation) between high/low patients for both initial function and subsequent recovery. Lastly, we examined the relative contribution of changes in brain structure and function to recovery in patients with low levels of initial function.

Low initial function and low subsequent recovery are related to lower sensorimotor β power and greater lesion-induced disconnection of contralateral [ipsilesional] white-matter motor projection connections. Moreover, differences in intra-hemispheric connectivity (structural and functional) are unique to initial motor function, while differences in inter-hemispheric connectivity (structural and functional) are unique to subsequent motor recovery.

Function-related and recovery-related differences in brain function and structure after stroke are related, yet not identical. Separating out the factors that contribute to each process is key to identifying potential therapeutic targets for improving outcomes.

## 1. Introduction

Stroke is a leading cause of disability worldwide. One in four people will have a stroke in their lifetime, and a quarter of those survivors remain moderately to severely disabled ten years later. Upper limb motor impairment is a common consequence of stroke that can limit activities of daily living and impact quality of life (Broeks et al., 1999).

Improving outcomes for stroke survivors will require an understanding of the mechanisms that support recovery. It is important to make the distinction between outcome and recovery. Outcome reflects the level of function/impairment at a given time post-stroke and is partially related to initial function/impairment. Recovery is a dynamic process defined as a return to or towards premorbid behavioural levels (Levin et al., 2009; Rothi & Horner, 1983). The fact that people with the same initial function/impairment can have different recovery profiles (Prabhakaran et al., 2008; Winters et al., 2015; Zarahn et al., 2011) indicates that the factors important for outcome and for subsequent recovery may be quite different (Ward, 2017). Identifying biomarkers of post-stroke recovery requires us to disentangle initial function/impairment-related and recovery-related differences in post-stroke brain function and structure. Functional and structural human neuroimaging provide complementary information to clinical measures (Laaksonen et al., 2012; Roiha et al., 2011; Ward, Brown, et al., 2006; Ward et al., 2003a) and can thus advance our understanding of recovery (Ward et al., 2003b, 2004; Ward, Newton, et al., 2006).

Previous investigations of the prognostic value of post-stroke brain structure have used lesion size, as well as the lesion’s direct and indirect effects on grey matter and white matter connections (Griffis et al., 2019, 2021; Rudrauf et al., 2008; Talozzi et al., 2023). Initial motor function/impairment and subsequent motor recovery show no or only weak relationships with lesion size (Alexander et al., 2010; C.-L. Chen et al., 2000; Egger et al., 2021; Zhu et al., 2010), but damage to key grey matter regions or white matter connections represent promising structural correlates of both initial motor function/impairment and subsequent motor recovery. The prognostic focus has largely been on the integrity of descending white matter projection connections, particularly the corticospinal tract (CST). CST integrity is related to post-stroke initial motor function/impairment (Maraka et al., 2014; Schulz et al., 2012; Swayne et al., 2008; Talelli et al., 2006) but can also account for differences in subsequent motor recovery independent of initial motor function/impairment (Byblow et al., 2015; Puig et al., 2017; Rapisarda et al., 1996; Stinear et al., 2007). Here we ask whether in addition to CST integrity, other structural connections (projection, commissural and association connections) are related to either initial motor function and/or subsequent motor recovery (Rondina et al., 2016, 2017).

Structural brain imaging cannot fully account for variability in either initial function/impairment or subsequent recovery. The additional prognostic relevance of brain function therefore becomes important, particularly those measures reflecting the early post-stroke balance between cortical inhibition and excitation that strongly influences the potential for experience dependent plasticity (Carmichael, 2012; Clarkson et al., 2010; Ward, 2017). Here we investigate the contribution of sensorimotor beta activity (β, ∼13-30Hz), which plays a vital role in the physiology and pathology of human movement and movement disorders. In fact, every movement is accompanied by a decrease in sensorimotor β activity (Event-Related Desynchronisation, suppression), which has been related to the activation of the sensorimotor cortex. The β suppression is followed by an increase in sensorimotor β activity (Event-Related Synchronisation, rebound), which has been related to active inhibition or the removal of excitation in the sensorimotor cortex (R. Chen & Hallett, 1999; Franzkowiak et al., 2010; Pfurtscheller, 1992; Salmelin et al., 1995). The β suppression-rebound complex is a robust phenomenon with high reproducibility (Espenhahn et al., 2017; Illman et al., 2021). Compared to healthy controls, stroke patients with upper limb impairments exhibit significantly lower β rebound in the acute and chronic phase providing a potential biomarker for motor function/impairment post stroke (Espenhahn et al., 2020; Laaksonen et al., 2012; Parkkonen et al., 2017, 2018; Tang et al., 2020). Changes in the β suppression-rebound complex during motor learning (Alayrangues et al., 2019; Haar & Faisal, 2020; Tan et al., 2014, 2016; Torrecillos et al., 2018) further strengthen the link between sensorimotor β activity and the experience dependent plasticity on which motor learning is based. Mechanistically, animal (Yamawaki et al., 2008) and human (Hall et al., 2010; Jensen et al., 2005) studies demonstrated that β activity is mediated by inhibitory interneuron drive via GABA-A receptors. Therefore, we focused on sensorimotor β activity assessed very early post-stroke (within 1 week) as a marker of the potential for experience dependent plasticity. We used tactile stimulation, which increases to β activity in the primary motor cortex (M1) and the primary and secondary sensory cortex (S1, S2), unconfounded by residual movement. In addition to M1 and S1, area S2 is of particular interest because of its anatomical connections to S1 and its functional role as an integration hub (Disbrow et al., 2003; Hinkley et al., 2007; Inoue et al., 2002; Krubitzer & Kaas, 1990; Lewis & Van Essen, 2000). Building upon previous work suggesting that modulatory afferent input may reach M1 via S2 (Laaksonen et al., 2012) we analyse the functional connectivity pattern of the three nodes: M1, S1, and S2.

Here we will address the following question: What are the key stroke-related changes in brain structure and brain function that are related to initial motor function and to subsequent motor recovery? We will then determine whether the process of recovery of motor function after stroke (independent of initial motor function) relies more on brain structure, brain function or both. Separating out the factors that contribute to initial motor function and those that are related to the subsequent motor recovery process itself is key to identifying potential therapeutic targets for promoting post-stroke motor recovery.

## 2. Materials and methods

Collecting high-quality neuroimaging data in acute stroke patients is extremely challenging, so here we capitalise on existing high-quality data. Data from two previously published articles, i.e. dataset 1 (Laaksonen et al., 2012) and dataset 2 (Parkkonen et al., 2018), were combined. Participants were recruited using the same inclusion and exclusion criteria, data were acquired in the same institute, and comparable experimental designs were used. We here conduct entirely new analyses to quantify post-stroke structural connectivity and functional connectivity between sensorimotor areas at the source level.

### 2.1 Experimental design

#### 2.1.1 Ethical approval

For both studies, the Local Ethics Committee of the Helsinki and Uusimaa Hospital District approved the study protocol, and all subjects provided written informed consent.

#### 2.1.2 Subjects

Patients with first-ever stroke in the middle cerebral artery territory causing unilateral upper limb impairments were recruited from the Department of Neurology, Helsinki University Hospital. Exclusion criteria were earlier neurological diseases, mental disorders, neurosurgical operations or head traumas, unstable cardiovascular/general condition. Eleven of the 18 patients used in (Laaksonen et al., 2012) were included, as 7 MRI scans were not available. From the 27 patients in (Parkkonen et al., 2018), 25 patients were used for this analysis, as 2 MRI scans were not available. Here we only include patients with MRI and MEG scans, thus, the total sample comprises 36 patients (18 females, age: *M* = 66.56; *SD* = 8.52; range 45-84 years; see **SI Table 1**).

#### 2.1.3 Time points and measurements

Data were recorded at three time points (**Fig. 1a**). For dataset 1 (Laaksonen et al., 2012), these time points are 1-7 days (T_0_), 1 month (T_1_), and 3 months (T_2_) post-stroke. For dataset 2 (Parkkonen et al., 2018), these time points are 1-7 days (T_0_), 1 month (T_1_), and 12 months (T_2_) post-stroke. Here we focus on the clinical data, MEG data and MRI data from T_0_ and use the clinical data from T_1_ for recovery-related analysis.

**Fig. 1.**
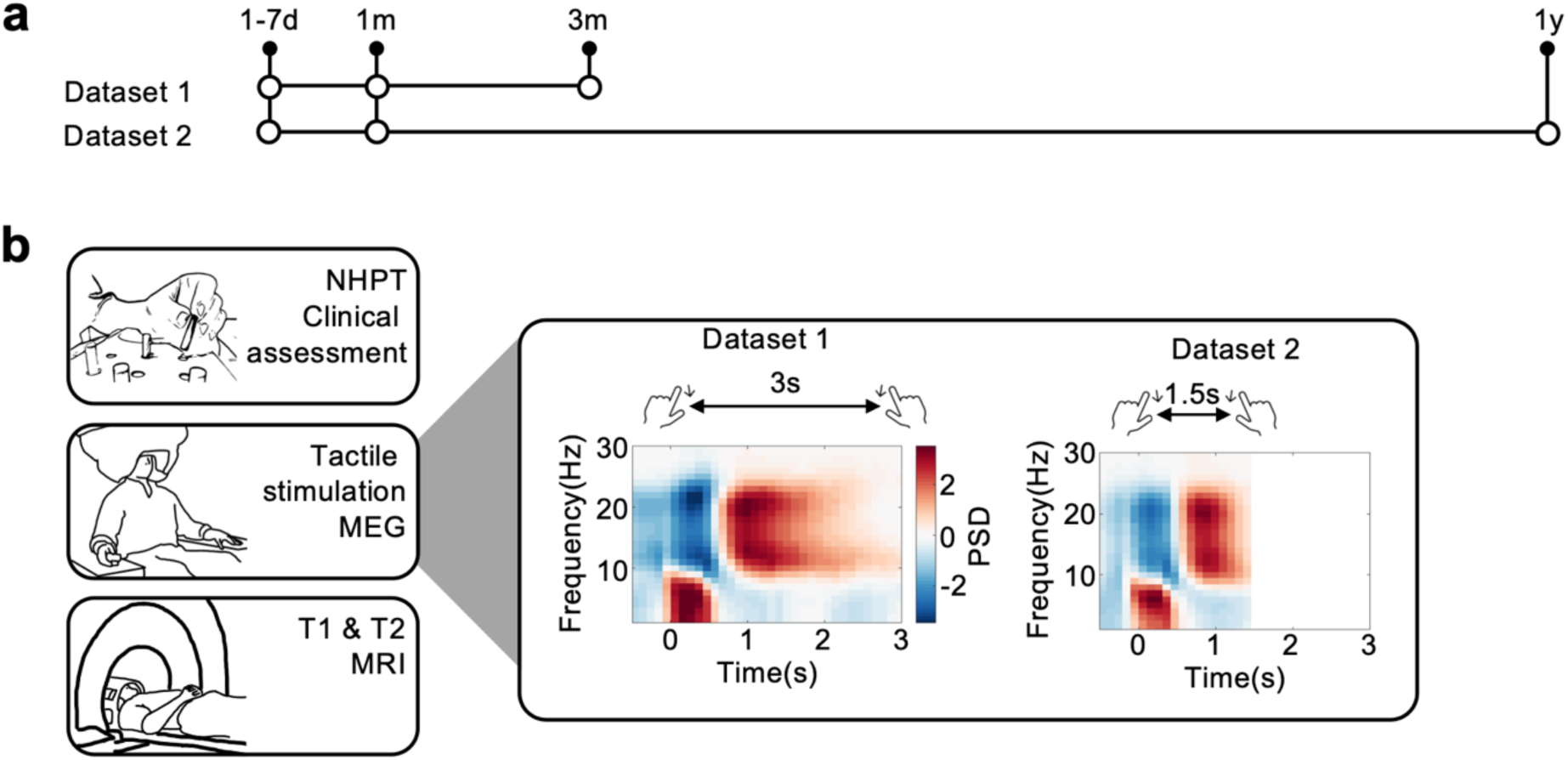
Study design. a) Timeline for dataset 1 (Laaksonen et al., 2012) and dataset 2 (Parkkonen et al., 2018), both comprising three assessment time points. b) Details of each assessment time point. Each assessment time point comprises a clinical assessment (NHPT) and MEG scan. In addition, a MRI scan was conducted at the first two assessment time points. MEG data were collected during tactile stimulation of the index finger. The interstimulus interval was 1.5s for dataset 2 and 3s for dataset 1. Power spectral density (PSD) is shown. In both datasets, clear β (13-30Hz) suppression (blue) and rebound (red) can be seen.

### 2.2 Clinical data

A series of clinical measures were obtained, see (Laaksonen et al., 2012; Parkkonen et al., 2018). Here we focus on manual dexterity quantified by the Nine-Hole Peg Test (NHPT). The NHPT demands well-functioning motor and somatosensory systems, as well as a fluent integration between these two. Therefore, it serves well as a clinical measure of upper limb motor function. Specifically, NHPT performance is quantified by the time taken to remove and replace nine pegs into nine holes, with a maximum time of 120 seconds in dataset 2 (Parkkonen et al., 2018), and 180 seconds in dataset 1 (Laaksonen et al., 2012). Based on the initial NHPT performance patients were grouped into low function patients (did not complete NHPT within 120 seconds) and high function patients (completed NHPT within 120 seconds, see **Fig. 2a**). Further, patients were grouped into patients who improved (difference between NHPT at T_1_ and T_0_ < 0) and patients who didn’t improve (difference between NHPT at T_1_ and T_0_ = 0, see **Fig. 5a**).

**Fig. 2.**
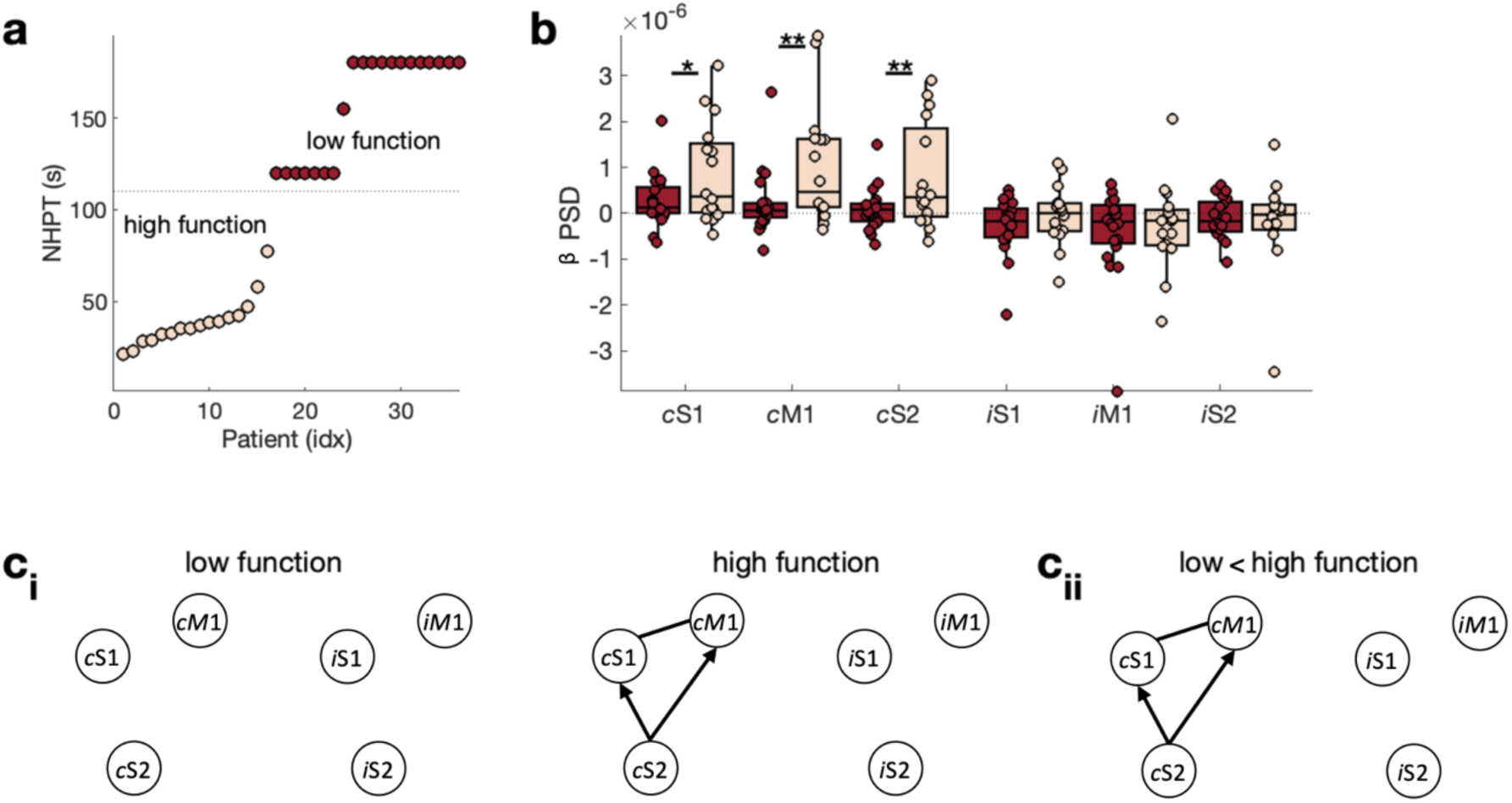
Behaviour and brain function for low and high function patients. a) Patients are grouped into high function and low function patients based on their NHPT performance at T_0_. b) Power (PSD) in the β frequency range (13-30Hz) during the β rebound for contralateral [ipsilesional] ‘c’ and ipsilateral [contralesional] ‘i’ M1, S1, and S2 ci) Connectivity strength and direction for low function (left) and high function (right) patients. Connections whose strength is significantly different from zero are highlighted by a solid line. Connections whose directionality is significantly different from zero have an arrow indicating the directionality. cii) Difference in connectivity strength and direction between low and high function patients.

### 2.3 MEG data

#### 2.3.1 MEG data acquisition

MEG data were acquired using a whole-scalp 306-channel MEG system (204 planar gradiometers and 102 magnetometers; Vectorview^TM^; Elekta Oy, Helsinki, Finland). Data were sampled at 944.8Hz with a band-pass filter of 0.03-308Hz in dataset 1 (Laaksonen et al., 2012), and at 1001.6Hz with a band-pass filter of 0.03-330Hz in dataset 2 (Parkkonen et al., 2018). Eye movements were simultaneously recorded via vertical electro-oculogram. Head position was recorded with respect to the MEG sensors using four head-position (HPI) coils. The locations of HPI coils, three anatomical fiducials (the nasion and two preauricular points) and head shape points (number of head shape points: *M* = 35.68; *SD* = 15.71; range = 11-61) were digitized using a 3D tracking system to allow alignment of the MEG and MRI coordinate system. Data were recorded according to the clinical condition of the patients, either in a sitting or supine position. A nurse inside the magnetically shielded room observed the patients for any possible movements.

#### 2.3.2 Tactile stimulation

MEG data were collected during tactile stimulation of the index finger. Tactile stimulation reliably induces a sensorimotor β suppression-rebound complex in healthy controls and stroke patients (Bardouille et al., 2010; Gaetz & Cheyne, 2006). Importantly, tactile stimulation targets purely tactile fibres and avoids inter- and intra-individual differences in movement ability, allowing direct cross-sectional and longitudinal comparisons.

Pneumatic diaphragms driven by compressed air were used to deliver tactile stimuli to the tip of the index finger. Stimuli were alternately delivered to both index fingers with an interstimulus interval of 3005ms in dataset 1 (Laaksonen et al., 2012) or 1500ms in dataset 2 (Parkkonen et al., 2018) (**Fig. 1b**). 60–80 stimuli were applied to each hand. The same stimulus intensity was applied to all subjects.

#### 2.3.3 MEG data pre-processing

A summary of the data processing pipeline is shown in **SI Fig. 1**. External noise was reduced from MEG data using the MNE-Python (version 0.22.0) implementation of temporal signal-space separation (tSSS)/Maxwell filtering. MEG pre-processing was performed using the Oxford Centre for Human Brain Activity (OHBA) Software Library (OSL, https://ohba-analysis.github.io/osl-docs/) version 2.2.0 using Matlab2022a. OSL builds upon Fieldtrip, SPM and FSL to provide a range of useful tools for M/EEG analyses. Continuous data were down-sampled to 250Hz and a band-pass filtered (1-45Hz). Time segments containing artefacts were identified using the generalised extreme studentized deviate method (GESD)(Rosner, 1983) on the standard deviation of the signal across all sensors in 1s non-overlapping windows, with a maximum number of outliers limited to 20% of the data and adopting a significance level of 0.05. Data segments identified as outliers were excluded from subsequent analyses. Further, denoising was conducted via independent component analysis (ICA) using temporal FastICA across the sensors (Hyvarinen, 1999). 62 independent components were estimated and components representing stereotypical artefacts such as eye blinks, eye movements, and electrical heartbeat activity were manually identified and regressed out of the data. Magnetometers and Planar-Gradiometers were normalised by computing the eigenvalue decomposition across sensors within each coil type and dividing the data by the smallest eigenvalue within each (Woolrich et al., 2011). Data were segmented from −0.5s to 1.5s or 3s depending on the interstimulus interval. Only trials with tactile stimulation of the affected hand were considered for this analysis. Registration between structural MRI and the MEG data was performed with RHINO (Registration of head shapes Including Nose in OSL), which makes an initial registration between the anatomical and polhemous fiducial landmarks. This fit is refined using an Iterative closest point (ICP) algorithm to optimise the correspondence between the polhemous headshape points and the mesh of the scalp extracted from the structural MRI. A single shell forward model was constructed using the individual inner skull mesh extracted from the structural MRI. Segmented data were projected onto an 8 mm grid in source space using a Linearly Constrained Minimum Variance (LCMV) vector beamformer (Van Veen & Buckley, 1988; Woolrich et al., 2011). Six regions of interest (ROIs) were considered: left and right M1, S1, and S2, as defined by the modified Schaefer Yeo parcellation ((Yeo et al., 2011), available from the *Lesion Quantification Toolkit* https://wustl.app.box.com/v/LesionQuantificationToolkit, (Griffis et al., 2021)). A single time course was estimated per ROI from the first principal component across the voxels within an ROI. Spatial leakage was attenuated using a symmetric multivariate leakage correction (Colclough et al., 2015, 2016).

#### 2.3.4 Functional Connectivity

MEG features were extracted in Python (version 3.9.9) with core dependencies as numpy (Harris et al., 2020) and scipy (Virtanen et al., 2020) using the Spectral Analysis In Linear Systems toolbox (Quinn & Hymers, 2020).

Linear dependencies between the six ROI time-series *X* were modelled using a multivariate autoregressive (MVAR) model of order *p* = 6 following the procedures outlined in (Quinn et al., 2021). For a review of these methods see (Blinowska, 2011).

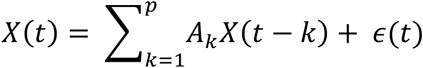

The autoregressive parameters *A* are transformed into the frequency domain using the Fourier transform:

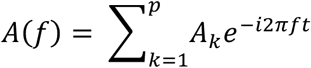

The spectral matrix *S(f)* is computed from *A(f)* and the residual covariate matrix 𝛴 The spectral matrix contains the power spectra of each region on the diagonal and the cross spectral densities on the off-diagonal elements.

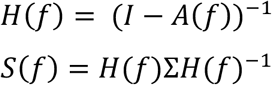

The spectral matrix is used to compute the power spectral density PSD(f) = S(f)/sample rate and the magnitude squared coherence (MSC).

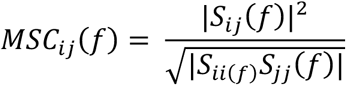

The MSC represents the cross-spectral density between two ROIs as a ratio of the power within each ROI. Finally, the Partial directed coherence (PDC) (Baccalá & Sameshima, 2001) is computed from the Fourier transform of the autoregressive parameters.

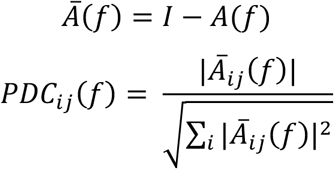

The PDC is closely related to the concept of Granger Causality. It ranges from zero to one and is normalised across columns of the inverse spectral matrix. The PDC at frequency *f* between signal *i* and *j* reflects the outflow of influence from *i* to *j* as a proportion of the total outflow from *i* to all nodes (Baccalá & Sameshima, 2001; Bastos & Schoffelen, 2016). In other words, if the PDC of the connection between *i* and *j* is large, it indicates that information in the recent past of time series *i* improves the prediction of the next step in time series *j* relative to how well the past of time series *i* improves prediction of all nodes.

In summary, PSD represents the strength of the beta activity within each ROI, whilst MSC represent the strength and PDC the direction of functional connectivity between ROIs. PSD, MSC, and PDC were baseline corrected (−0.5s to 0s) and subsequently averaged across the β rebound time window (0.6s to 1.2s).

### 2.4 MRI data

#### 2.4.1 MRI data acquisition

The MRI protocol was acquired using a 3T Philips Achieva MRI scanner (Philips Medical Systems, The Netherlands). A high-resolution 3D T1-weighted scan (T1 3D TFE SENSE, TR = 9.9 ms, TE = 4.6 ms, voxel size = 0.88 × 0.83 × 0.83 mm^3^, dimensions = 187 × 288 × 288 slices, flip angle = 8 degrees, bandwidth = 149) and a 3D T2-weighted scan was acquired (T2 TSE 4mm CLEAR, TR = 4000 ms, TE = 80 ms, voxel size = 0.469 × 0.469 × 4.4 mm^3^, dimensions = 512 × 512 × 32, flip angle = 90 degrees, bandwidth = 216).

#### 2.4.2 Lesion mapping

Stroke lesions were demarcated using the semi-automated segmentation algorithm *Clusterize* (https://www.medizin.uni-tuebingen.de/de/das-klinikum/einrichtungen/kliniken/kinderklinik/kinderheilkunde-iii/forschung-iii/software) applied to the axial T2 image acquired at T_0_. Agreement between a manual segmentation and the semi-automated lesion maps obtained with *Clusterize* has been shown to be excellent in acute stroke using CT, DWI and T2 FLAIR (de Haan et al., 2015; Ito et al., 2019; Wilke et al., 2011). The resulting lesions were manually verified and if necessary corrected. Lesion maps were smoothed using a 2 mm full-width half maximum (FWHM) Gaussian kernel. Lesions were normalised to standard MNI space and left hemispheric lesions were flipped.

#### 2.4.3 Lesion size and ROI damage

To investigate if differences in initial motor function and subsequent recovery are due to lesion size or direct damage to M1, S1 or S2 cortex, we calculated the lesion size and the ROI damage for M1, S1, and S2 using *Lesion Quantification Toolkit* (https://wustl.app.box.com/v/LesionQuantificationToolkit)(Griffis et al., 2021). Specifically, ROI damage quantifies the overlap between the lesion and each ROI as a percent of voxels with the ROI that overlaps with the lesion. Despite some limitations (Seghier, 2023), ROI damage can provide a straightforward way of reducing the dimensionality of the lesion’s effect on brain structure.

#### 2.4.4 Structural connectivity

To characterise structural connections as well as the relationship between functional and structural connections, we quantified voxel-wise percent disconnection maps and the effect of the lesion on the relevant association, projection, and commissural connections, again using the *Lesion Quantification Toolkit* (Griffis et al., 2021).

The voxel-wise percent disconnection map indicates for each voxel the percentage of all the streamlines (computed from the HCP-842 streamline tractography atlas) in that voxel relative to those streamlines that are expected to be disconnected by the lesion (for more details see (Griffis et al., 2021)).

For projection connections and commissural connections, lesion-related damage to white matter tracts (i.e., tract disconnection) was quantified. Tract disconnection is the percent of streamlines of the HCP-842 population-average streamline tractography atlas that intersect the lesion (for more details see (Griffis et al., 2021)). While the atlas comprises 70 tracts here we focus on the motor projection connections (corticospinal tract [CST], corticostriatal pathway [CS], corticothalamic pathway [CT], frontopontine tract [FPT], parietopontine tract [PPT]) and motor commissural connections (mid-anterior corpus callosum, central corpus callosum, mid-posterior corpus callosum).

The structural connection between the cortical ROIs M1, S1, and S2, i.e., association connections, cannot simply be assessed using the 70 tracts of the HCP-842 population-average streamline tractography atlas. Therefore, these association connections were quantified using the structural shortest path lengths (SSPL) between M1, S1, and S2. SSPL reflects the minimum number of direct parcel-to-parcel (using the modified Schaefer Yeo parcellation ((Yeo et al., 2011)) white matter connections (computed from the HCP-842 streamline tractography atlas) that must be traversed to establish a structural path between two ROIs (for more details see (Griffis et al., 2021)). Here we report the lesion-induced increases in SSPLs relative to the atlas SSPL matrix.

### 2.5 Statistical analysis

Null-hypothesis testing was carried out with non-parametric permutations (Maris & Oostenveld, 2007; Nichols & Holmes, 2002). Depending on the hypothesis test, different forms of non-parametric permutation are used, though the overall procedure is similar. To compare differences between subgroups row-shuffle permutation is used. To test whether a measurement deviates from zero sign-flipping permutation is used. A null distribution of the test statistic is derived by recomputing the test statistic after each permutation. The observed test statistic is then compared to this null distribution and is ‘significant’ if it exceeds a pre-set critical threshold. Here we build the null distribution from 5000 repetitions and use the 95^th^ percentile (indicated with *) and the 99^th^ percentile (indicated with **) of the null distribution as thresholds.

For prediction analysis forward stepwise linear regression was used to identify possible predictors of the outcome improvement status (improved = 0, didn’t improve = 1) as implemented in R (version 4.0.2). Predictors were standardised using z-transformation. At each step, predictors were included when *p*<0.15 (Wald test) and removed when *p*>=0.15 (Wald test). Predictors showing high collinearity (variance inflation factor (VIF) > 2.5) were re-assessed. A backward stepwise approach was used to test the stability of the model (inclusion criterion: *p*>= 0.15; removal criterion: *p*<0.15; Wald test). The Brier score and area under the curve (AUC) of the receiver operating characteristic curve (ROC) were used to quantify the goodness of fit of the logistic regression model. Finally, accuracy, sensitivity, specificity, positive predictive value (PPV), and negative predictive value (NPV), including the corresponding 95% CIs, were calculated using two-way contingency tables.

### 2.6 Data availability

We will consider requests to access the data in a trusted research environment as part of a collaboration if requirements of EU data protection and Finnish legislation on health data are followed. Contact.

## 3. Results

### 3.1 Acute post-stroke brain function and structure relate to initial motor function

First, we assess motor function-related differences in brain function and structure in the acute phase post-stroke (T_0_). To this end, patients are grouped based on their initial NHPT scores into low and high function patients. Patients who could not complete the NHPT at T_0_ within 120 seconds are referred to as low function patients (**Fig. 2a**; N = 20; 11 females, age: *M* = 67.85; *SD* = 9.11; range 47-84 years), while patients who could complete the NHPT within 120 seconds are referred to as high function patients (N = 16; 7 females, age: *M* = 64.94; *SD* = 7.70; range 45-76 years). Based on these definitions, patients fall into two distinct groups (**Fig. 2a**).

#### 3.1.1 Brain function

We then asked if brain function differs between high and low function patients. Extending the previous reports on the MEG sensor level (Laaksonen et al., 2012; Parkkonen et al., 2018), we quantified the sensorimotor β rebound on source level from contralateral [ipsilesional] and ipsilateral [contralesional] sensorimotor ROIs (M1, S1, S2). In line with the sensor level analyses, we found that the sensorimotor β rebound was significantly reduced in the contralateral [ipsilesional] M1 (*t(34)*=-2.39, *p*<0.05), S1 (*t(34)*=-2.10 *p*<0.01), and S2 (*t(34)*=-2.48, *p*<0.01) in initially low function patients when compared to high function patients (**Fig. 2b**). No significant differences were observed for the homologous ipsilateral [contralesional] ROIs (all three *p’s*>0.05).

To further understand the underlying mechanisms of these motor function-related differences in the sensorimotor β rebound, we next investigated the strength (i.e., Magnitude-Squared Coherence, MSC) and directionality (i.e., Partial Directed Coherence, PDC) of functional connectivity. Regarding intra-hemispheric functional connectivity strength, in low function patients none of the connections were strong enough to reach statistical significance (**Fig. 2c_i_**, **SI Fig. 2ai**). In contrast, in high function patients all contralateral [ipsilesional] intra-hemispheric functional connections were strong enough to pass the significance threshold (S1-M1 [*t(15)*=2.73, *p*<0.01], S2-S1 [*t(15)*=2.04, *p*<0.05], S2-M1 [*t(15)*=2.57, *p*<0.01]), resulting in significant differences in functional connectivity strengths between high and low function patients (all three *p*’s<0.05, **Fig. 2c_ii_**, **SI Fig. 2aii**). Inter-hemispheric connections were not significant in either patient group. Analysing the directionality of functional connectivity showed no significant directionality (i.e., none of the connections showed a directionality that was significantly different from zero) in low function patients. In contrast, in high function patients contralateral [ipsilesional] S2-M1 (*t(15)*=-2.34, *p*<0.05) and S2-S1 (*t(15)*=-3.45, *p*<0.01), were driven by S2 (**Fig. 2c_i_**, **SI Fig. 2b^i^)**, and the directionality of these functional connections differs significantly between the high and low function patients (both *p’s*<0.05, **Fig. 2c_ii_**, **SI Fig. 2b^i^**).

#### 3.1.2 Brain structure

Next, we asked if brain structure differs between high and low function patients. Lesion volume, as well as extent of M1, S1, and S2 damage, did not significantly differ between low and high function patients (all *p’s*>0.05, **Fig. 3a,b**), indicating that the differences in initial motor function patients are not simply explained by these direct lesion characteristics. We complemented our functional connectivity analysis with structural connectivity analysis. Descriptively, low function patients show higher overall tract disconnection than high function patients (**Fig. 4a**). Statistically, we observed significant differences in the association connections between S2-S1 (*t(34)*=1.85, *p*<0.05) and S2-M1 (*t(34)*=2.67, *p*<0.01), with longer structural shortest path lengths (SSPLs), for low function patients (**Fig. 4b**). Similarly, motor projection connections (i.e., CST, CS, CT, FPT, PPT) were significantly different between groups (*t(34)*=3.04, *p*<0.01, **Fig. 4b**, for individual tracts see **SI Fig. 3a**), with higher tract disconnect for low function patients, while the inter-hemispheric commissural connections (i.e., Mid Anterior, Central, Mid Posterior commissural connections) did not differ significantly between low and high function patients (*p*>0.05, **Fig. 4b**, **SI Fig. 3b**).

**Fig. 3.**
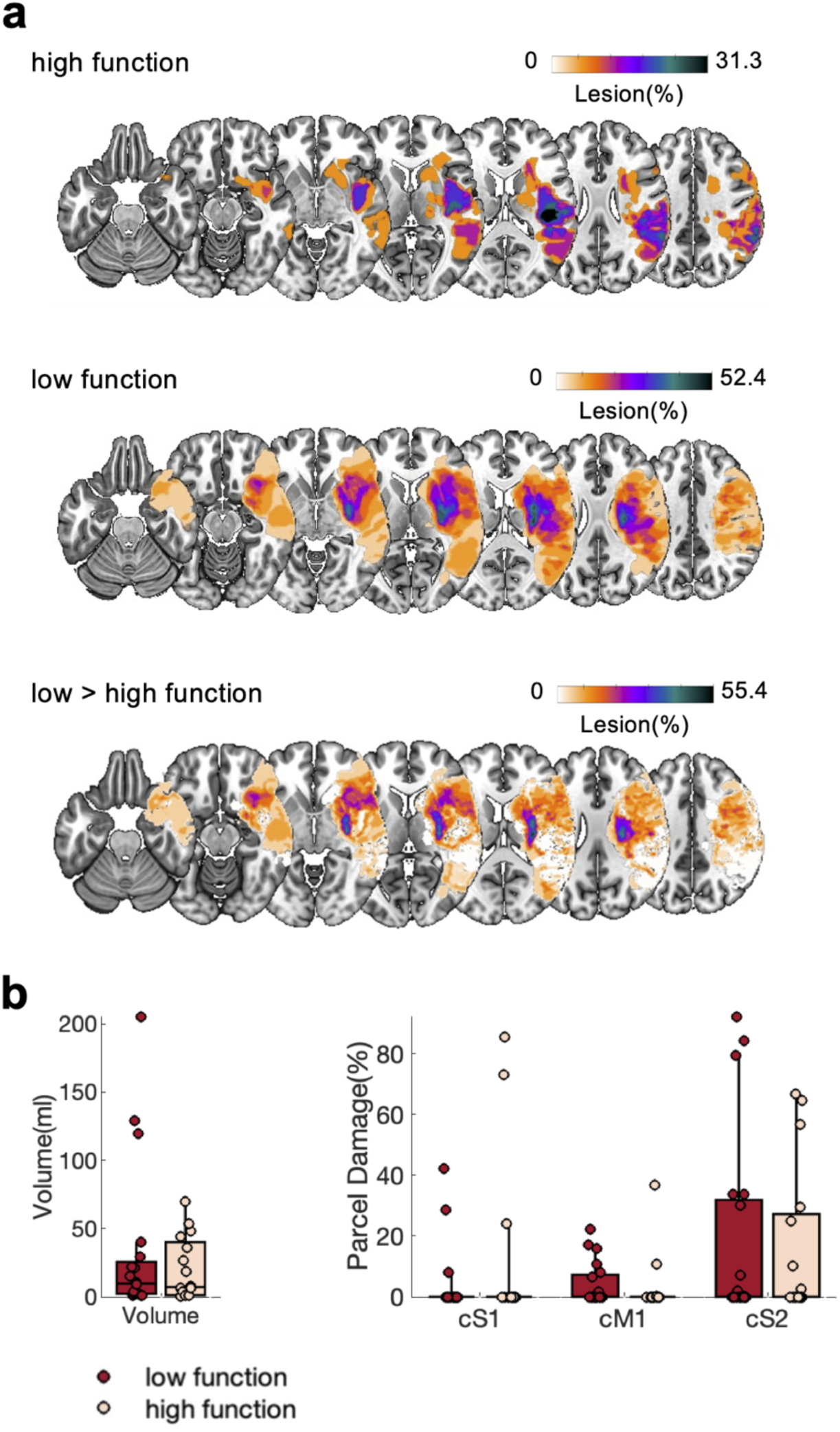
Lesion maps, lesion volume, and S1, M1, S2 ROI damage for low and high function patients. a) Heatmap of lesions for high function patients, low function patients, and the difference map between low and high function patients. Left hemispheric lesions were flipped. Heatmaps are overlaid on an MNI template. b) Lesion volume (left) and ROI damage for contralateral (ipsilesional) S1, M1, and S2.

**Fig. 4.**
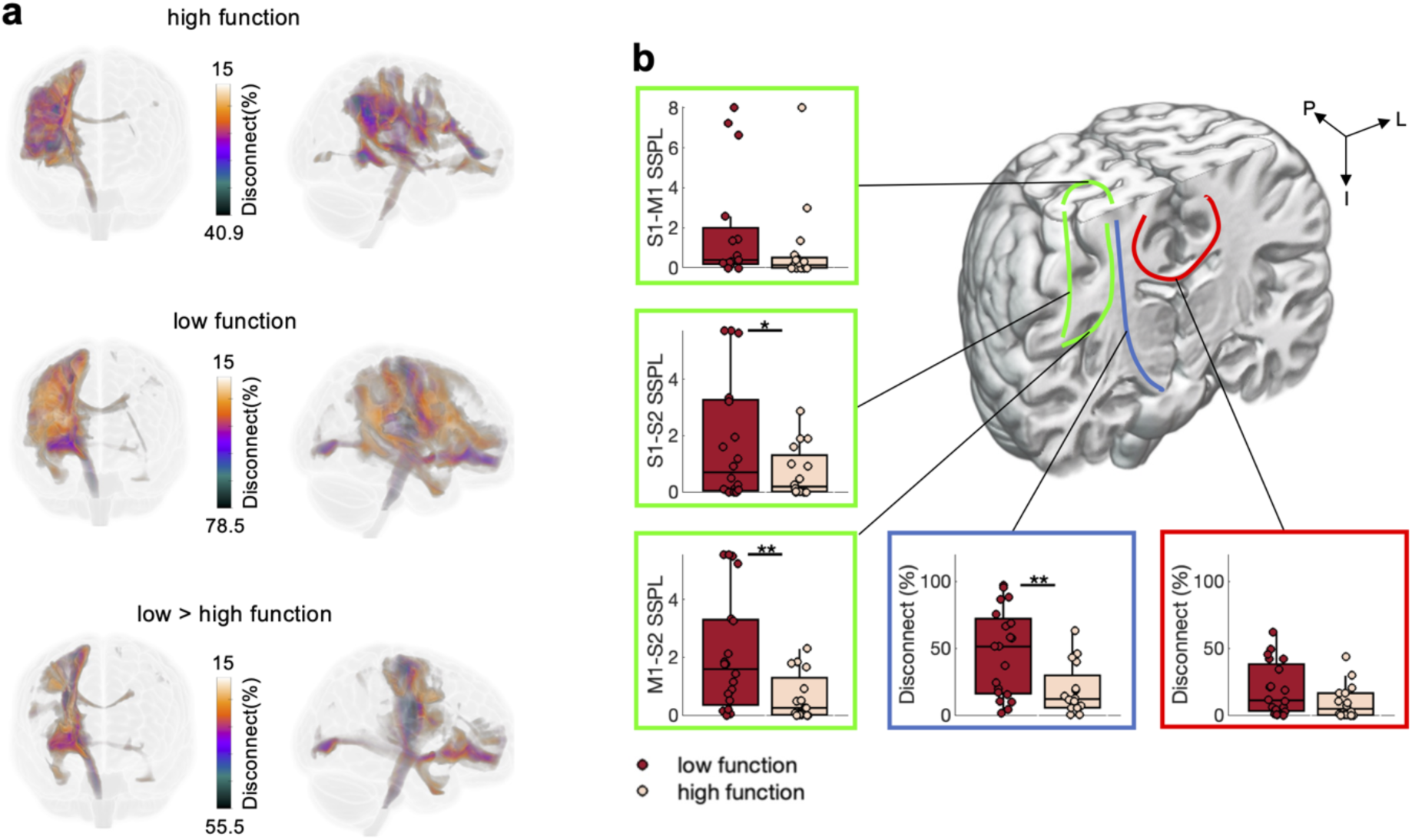
Structural connectivity for low and high function patients. a) Voxel-wise percent disconnection maps from the frontal and lateral view. b) Association connections (green) quantified using structural shortest path lengths (SSPL) between M1, S1, and S2. The average across motor projection connections (blue, see **SI Fig. 3a** for individual motor projection connections) and the average across commissural motor connections (red, see **SI Fig. 3b** for individual commissural motor connections). Note that the highlighted connections on the coronal slice are only schematic representations. Significant differences between groups are highlighted (p<0.05 *, p<0.01**).

To summarize, initial motor function, as quantified by the NHPT, is related to brain function and brain structure. At the level of brain function, low initial motor function is related to a lower β rebound in contralateral [ipsilesional] M1, S1, and S2, as well as lower functional connectivity strength and directionality between these areas. At the level of brain structure, low initial motor function is related to less direct association connections between these areas and by higher disconnection of projection connections.

### 3.2 Acute post-stroke brain function and structure relate to subsequent motor recovery

Next, we sought to explore whether brain function and structure in the acute stage (T_0_) can help distinguish between patients who subsequently recover from patients who subsequently don’t recover (T_1_). As subsequent motor recovery is strongly related to initial motor function/impairment (Prabhakaran et al., 2008; Winters et al., 2015) investigating purely recovery-related processes requires careful correction for initial function/impairment or a group of patients with similar initial function/impairment yet different subsequent recovery trajectories. Here we focus on the group of initially low function patients, defined as those patients who could not complete the NHPT at T_0_ within 120 seconds (**Fig. 2a**). These patients were then divided based on the change in their NHPT performance (NHPT T_1_ – NHPT T_0_) over 1 month, using < 0 as the cut-off. Of the initially low function patients, eight patients improved in the NHPT (3 females, age: *M* = 69.63; *SD* = 9.21; range 57-84 years), while twelve patients didn’t improve (8 females, age: *M* = 66.67; *SD* = 9.25; range 47-78 years) (**Fig. 5a**).

**Fig. 5.**
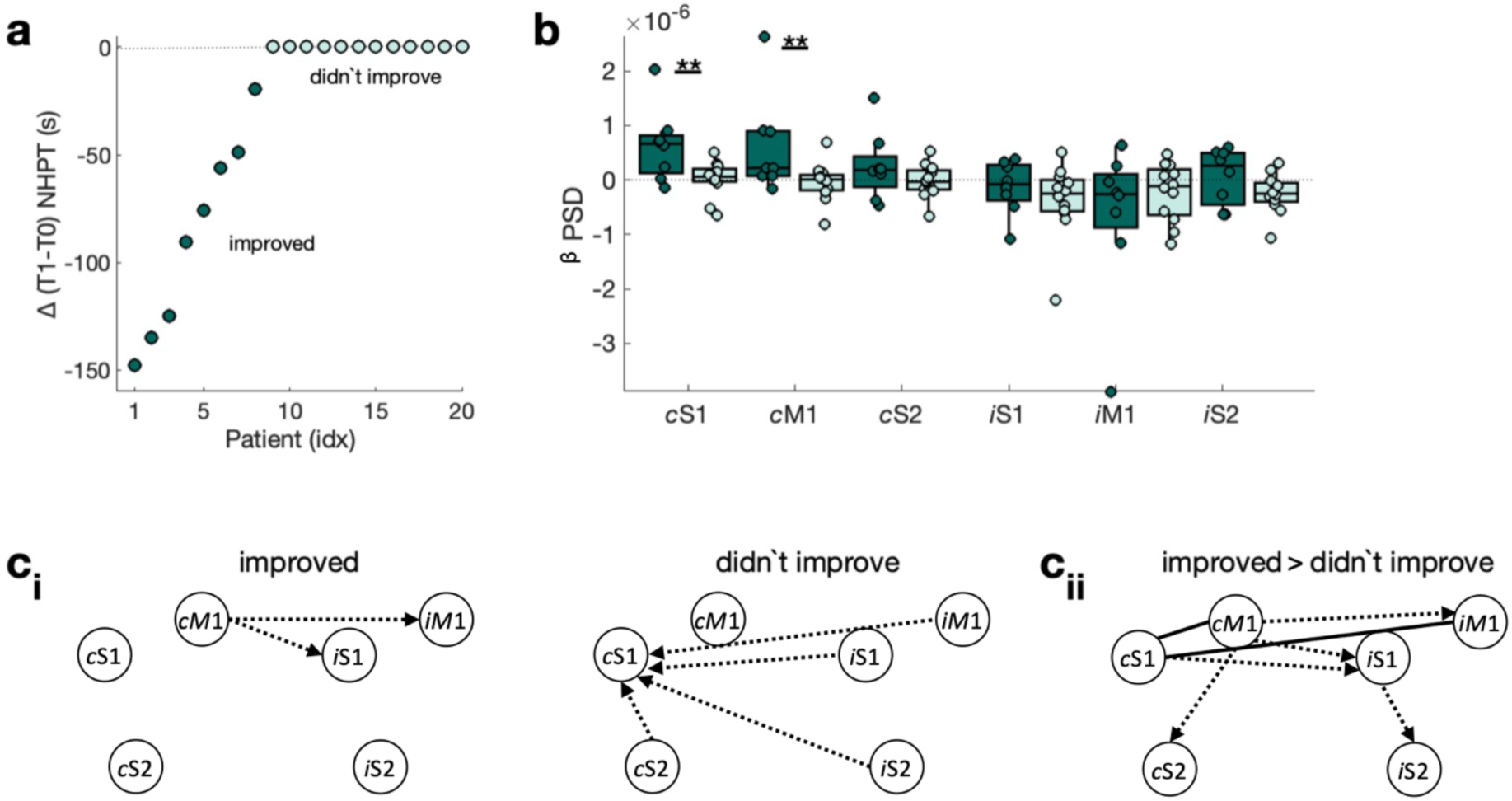
Behaviour and brain function for patients who improved and patients who didn’t improve considering only patients who have initially low motor function. a) Patients are grouped into ‘improved’ and ‘didn’t improve’ based on their difference in NHPT performance (i.e., T_1_ −T_0_). b) Power (PSD) in the β frequency range (13-30Hz) during the β rebound for contralateral [ipsilesional] ‘c’ and ipsilateral [contralesional] ‘i’ M1, S1, and S2 ci) Connectivity strength and direction for patients who improve (left) and patients who didn’t improve (right). Connections whose strength is significantly different from zero are highlighted by a solid line. Connections whose directionality is significantly different from zero have an arrow indicating the directionality. If only the direction, but not the strength is significant the connection is shown as a dashed line. cii) Difference in connectivity strength and direction between patients who improve and patients who didn’t improve.

#### 3.2.1 Brain function

First, we asked if brain function differs between patients who improved and patients who didn’t improve. Patients who improved from T_0_ to T_1_ have significantly higher sensorimotor β rebound in the contralateral [ipsilesional] M1 (*t(18)*=2.26, *p*<0.01) and S1 (*t(18)*=2.77, *p*<0.01) at T_0_ (**Fig. 5b**). Further, analysis of functional connectivity strength (i.e., MSC) showed no significant connections in either group. Subsequent analysis of functional connectivity direction (i.e., PDC) revealed that inter-hemispheric functional connectivity at T_0_ is driven by the contralateral [ipsilesional] hemisphere in patients who improve from T_0_ to T_1_ (**Fig. 5c_i_**, **SI Fig. 4b_i_**, **SI Results**), while patients who didn’t improve from T_0_ to T_1_ exhibit the opposite directionality (i.e., driven by the ipsilateral [contralesional] hemisphere, **Fig. 5c_i_**, **SI Fig. 4b_i_**, **SI Results**), resulting in significant differences between patients who improve and patients who didn’t improve (**Fig. 5c_ii_**, **SI Fig. 4b_ii_**, **SI Results**).

#### 3.2.2 Brain structure

Next, we asked if brain structure differs between patients who improved and patients who didn’t improve. Patients who didn’t improve from T_0_ to T_1_ have larger lesions at T_0_ (*t(18)*=2.01, *p*<0.05, **Fig. 6b**), but no significant differences in ROI damage were observed (all three *p’s*>0.05, **Fig. 6b**). Structural connectivity analysis further revealed no significant group differences for association connections between M1, S1, and S2 (*p’s*>0.05, **Fig. 7b**) at T_0_. However, both, motor projection connections (i.e., CST, CS, CT, FPT, PPT, *t(18)*=-2.34, *p*<0.05, **Fig. 7b**, for individual tracts see **SI Fig. 5a**) and motor commissural connections (i.e., Mid Anterior, Central, Mid Posterior commissural connections, *t(18)*=-3.08, *p*<0.01, **Fig. 7b**, for individual tracts see **SI Fig. 5b**) at T_0_, were significantly different between groups (both *p’s*<0.01), with higher tract disconnect for patients who didn’t improve from T_0_ to T_1_.

**Fig. 6.**
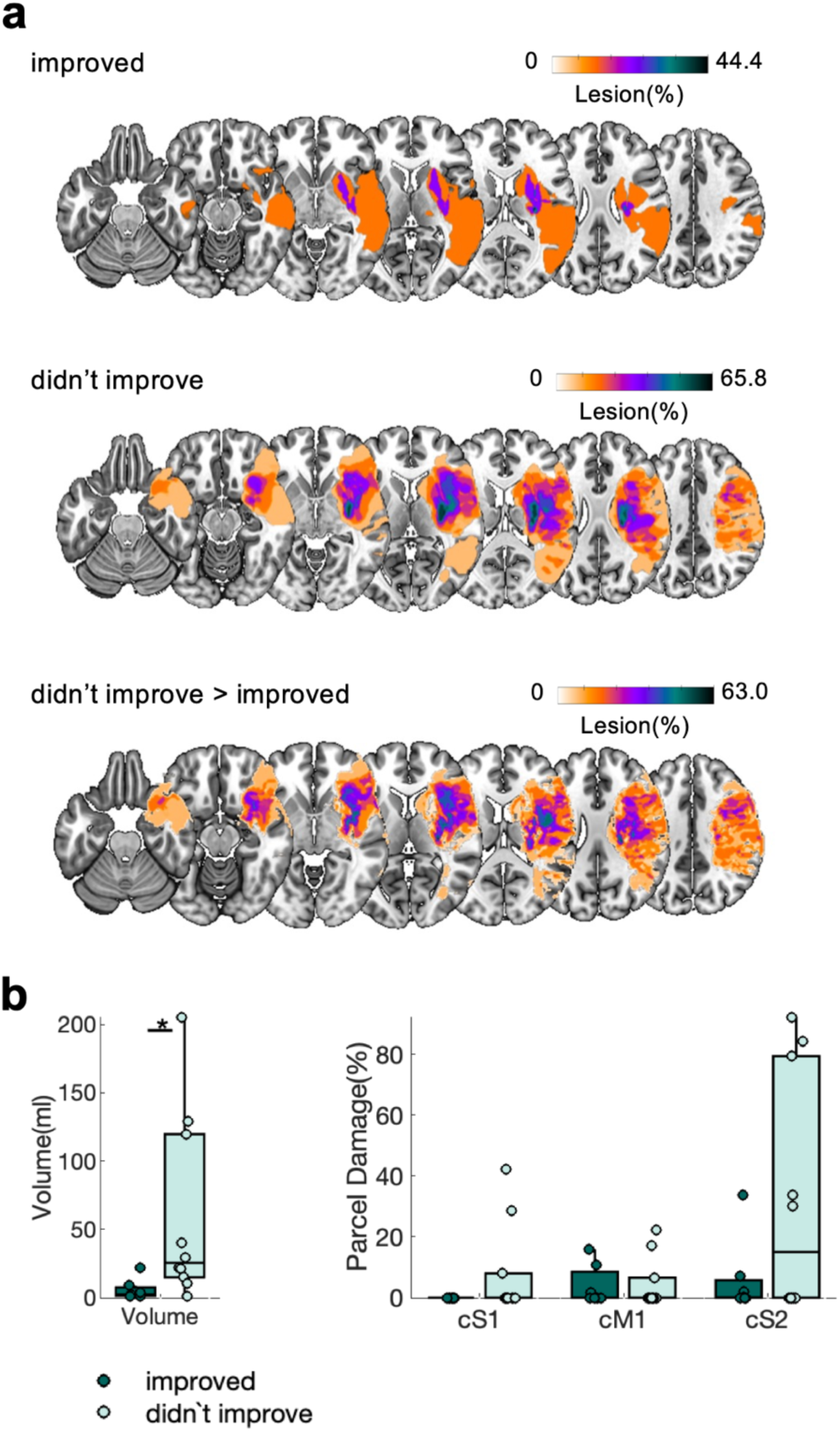
Lesion maps, lesion volume, and S1, M1, S2 ROI damage for patients who improved and patients who didn’t improve considering only patients who have initially low motor function. a, b) Same as Fig. 3. p<0.05 *.

**Fig. 7.**
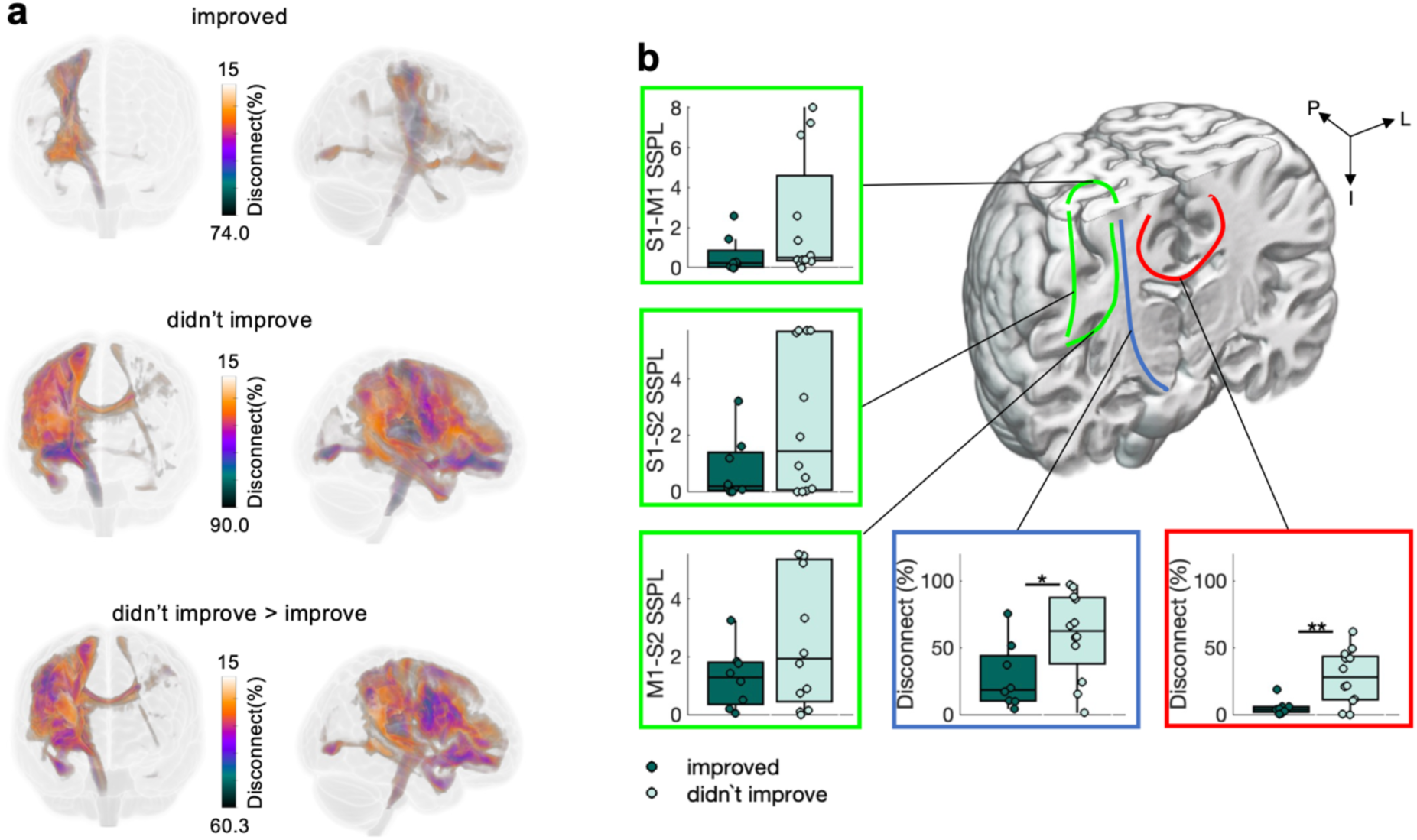
Structural connectivity for patients who improve and patients who didn’t improve considering only patients who have initially low motor function. a, b) Same as Fig. 4. See **SI Fig. 5a,b** for more details.

Together, subsequent motor recovery in patients with initially low motor function, as quantified by improvement in the NHPT from T_0_ to T_1_, relates to brain function and brain structure acquired at T_0_. At the level of brain function, severely affected patients who improve motor function show a stronger T_0_ β rebound in contralateral [ipsilesional] M1 and S1, and T_0_ inter-hemispheric connectivity driven by the contralateral [ipsilesional] hemisphere. At the level of brain structure, severely affected patients who improve motor function have smaller lesions and more intact motor projection and motor commissural connections.

### 3.3 Predicting subsequent motor recovery in stroke patients who have initially low motor function using multimodal neuroimaging

Finally, we explored the potential to predict subsequent motor recovery in stroke patients who have initially low motor function using multimodal neuroimaging. We focus on stroke patients who have initially low motor function as initial motor function/impairment alone is insufficient to reliably predict subsequent recovery in this subsample of patients (Prabhakaran et al., 2008; Winters et al., 2015). To avoid overfitting, we focussed on the brain functional and structural properties at T_0_ which showed a significant difference between initially low function patients who improved and initially low function patients who didn’t improve from T_0_ to T_1_ (see Section 3.2). To recap, at the level of brain function these properties are M1 β rebound (see **Fig. 5b**), S1 β rebound (see **Fig. 5b**), strength of intra-hemispheric S1-M1 connectivity (MSC) (see **Fig. 5c_ii_**), and strength of inter-hemispheric S1-M1 connectivity (MSC) (see **Fig. 5c_ii_**). At the level of brain structure these properties are lesion size (see **Fig. 6b**), motor projection connections (see **Fig. 7b**), and motor commissural connections (see **Fig. 7b**).

From these candidate variables measured at T_0_, leave-one-out cross-validation and a forward stepwise logistic regression were used to identify possible predictors of subsequent motor recovery from T_0_ to T_1_. Strong collinearity was found between M1 and S1 β rebound, and so these variables were consequently averaged. The forward stepwise logistic regression reduced the significant predictors to (i) strength of inter-hemispheric S1-M1 connectivity (MSC) (β = −2.42, 95% CI = −4.76-0.10, *p* = 0.042) and (ii) integrity of motor projection connections (β = 1.46, 95% CI = −0.18-3.09, *p* = 0.080). The Brier score (0.10) and the AUC of the ROC (0.92) suggest an excellent fit (for more model fit measures and alternative models see **SI Results**). Initially low function patients who had stronger functional inter-hemispheric S1-M1 connectivity and more intact motor projection connections at T_0_ were likely to show subsequent motor recovery (**SI Fig. 6**). The accuracy of the model was 0.90 (95% CI = 0.68-0.99), the sensitivity 0.91 (95% CI = 0.62-0.99), and the specificity was 0.88 (95% CI = 0.47-0.99), whereas the PPV and NPV were, respectively, 0.92 (95% CI = 0.64-0.99) and 0.88 (95% CI = 0.51-0.98). These results were confirmed by forward stepwise analysis.

## 4. Discussion

Here we asked what differences in acute post-stroke brain structure and function can account for differences in initial motor function and subsequent recovery of motor performance. To address these questions, we capitalised on hard-to-come-by high-quality MEG and MRI data collected in the first week post-stroke (Laaksonen et al., 2012; Parkkonen et al., 2018). We found that low initial motor function and low subsequent motor recovery are related to lower sensorimotor β rebound and greater lesion-induced disconnection of contralateral [ipsilesional] white-matter motor projection connections. Moreover, unique to initial motor function are differences in functional and structural intra-hemispheric connectivity, while differences in functional and structural inter-hemispheric connectivity are unique to subsequent motor recovery.

### 4.1 β rebound and projection connections as functional and structural markers for initial motor function and subsequent motor recovery

In the first week post-stroke β rebound was lower (i) in low compared to high function patients; and (ii) in initially low function patients who improve motor function compared to patients who didn’t improve motor function. This is in line with previous studies showing that β rebound correlates with initial motor function/impairment (i.e., patients with lower β rebound show low function/high impairment) and subsequent motor recovery (i.e., patients with lower β rebound show low subsequent recovery) (Laaksonen et al., 2012; Parkkonen et al., 2017, 2018; Tang et al., 2020). Collectively these findings suggest that early post-stroke β rebound is a functional marker for initial motor function/impairment and subsequent motor recovery. Overall, our MEG findings are in keeping with work in pre-clinical models of stroke that report early reduced neuronal activity in peri-infarct regions followed by restoration of activity (both peri-infarct and network connectivity) associated with recovery of function (see (Campos et al., 2023) for review). More specifically, lower β rebound is linked to lower GABA levels (Gaetz et al., 2011) and higher M1 cortical excitability (Hari et al., 1998; Salenius et al., 1997; Salmelin & Hari, 1994), so our results suggest that in the low function and poorer recovering patients, there is some very early motor cortex hyperexcitability, in keeping with previously observed reduced short interval cortical inhibition (SICI, related to GABA_A_ signalling) and shlong interval cortical inhibition (LICI, related to GABA_B_ signalling) in acute stroke. Several techniques have been used to assess markers of cortical excitability after stroke in humans (Mäkelä et al., 2015; Motolese et al., 2023), which point to increased cortical excitability (or reduced inhibition) in the acute phase post stroke. This cortical hyperexcitability has often been highlighted as something that supports recovery (in animal models) by enhancing the potential for experience-dependent plasticity (Carmichael, 2012; Ward, 2017), but in our patients, cortical hyperexcitability does not appear to be beneficial.

The explanation for this might come in the changes in brain structure, where we demonstrated that, like β rebound, higher disconnection of several white matter projection connections is related to low initial motor function and low subsequent motor recovery. One idea is that the observed changes in cortical excitability (and by extension enhanced plasticity) cannot exert beneficial effects over motor recovery because of the disconnection of projections to spinal cord motoneurons or contralesional hemisphere. This hypothesis will need to be addressed in future studies.

### 4.2 Cortical intra-hemispheric connectivity differs as a function of initial motor function

To further understand the function- and recovery-related reduction in the sensorimotor β rebound we asked if intra- and inter-hemispheric differences in functional and structural connectivity relate to initial motor function and subsequent motor recovery. Intra-hemispherically, we observed function-related, but not recovery-related, differences in functional and structural connectivity between primary and secondary somatosensory areas and between motor and somatosensory areas. Specifically, we found that the ipsilesional M1-S1 connection was stronger in high function patients and patients who recovered. In line with previous work, we found no clear directionality between M1 and S1 (Gandolla et al., 2021). While some studies provide clear evidence for M1 influencing S1, for example, the neural activity in somatosensory areas is modified by motor tasks (Ageranioti-Bélanger & Chapman, 1992) or evidence from animal studies suggests that M1 provides weak input to nearly all pyramidal neurons in S1 (Kinnischtzke et al., 2016). Other studies showed that input to S1 influences M1. For example, sensory stimulation facilitates functional reorganization of M1 (Garry et al., 2005; Hamdy et al., 1998; Ridding & Ziemann, 2010), Transcranial Magnetic Stimulation over S1 increases M1 excitability in healthy individuals (de Freitas Zanona et al., 2023) and motor learning post-stroke (Brodie et al., 2014), and animal data suggest that S1 input to M1 pyramidal cells can drive postsynaptic activity (Petrof et al., 2015). Together, it seems that ipsilesional M1-S1 are strongly and reciprocally connected to ensure precise movements, and deficits in this connection are related to low motor function.

Regarding the connections between M1 and S2 as well as S1 and S2, we observed structurally a longer path length for low function patients than for high function patients, indicating that lesions disrupted the shortest path in low function patients. In addition, on the functional level, we found significant strength and directionality in functional connectivity in high function patients, with M1 and S1 both being driven by S2, whereas in low function patients no functional connectivity was found between these areas. S2 is the first cortical area that unites sensory information from the two body halves, thus it has been thought to be an important area for sensorimotor integration (Hinkley et al., 2007; Inoue et al., 2002) and bimanual tasks (Disbrow et al., 2001). This explains its dense connection with several areas in the parietal and frontal cortex, such as the posterior parietal and premotor areas (Disbrow et al., 2003; Krubitzer & Kaas, 1990; Lewis & Van Essen, 2000). Here, we observed a directionality whereby S2 was the leading area. This might be unexpected, given that both, S2 and S1, receive direct input from the ventroposterior thalamus (for review see (Jones, 1985), (Disbrow et al., 2002; Friedman & Murray, 1986)), which informed the theory that cortical somatosensory processing depends on hierarchically equivalent and parallel processing in S1 and S2 (Mountcastle, 1978, 1986; Rowe et al., 1996). The directionality of the interaction between S1 and S2 has been probed using selective inactivation in marmosets, revealing that ∼70% of S2 neurons showed reduced activity when S1 was inactivated (Zhang et al., 1996), while 35% of S1 neurons showed reduced activity when S2 was inactivated (Zhang et al., 2001). This asymmetry could be due to anatomical asymmetries (for review see (Burton, 1986; Jones, 1986)), which led to the hypothesis that the S1 to S2 input represents a feed-forward projection, whereas the S2 to S1 input is a feed-back projection (Jones, 1986).

Together, while contralateral [ipsilesional] sensorimotor β rebound is reduced in low function patients and in patients who didn’t recover, differences in functional and structural intra-hemispheric connectivity are unique to the level of motor function. Thus, one could argue that sensorimotor β rebound and good manual dexterity demand sufficient structural and functional integrity between the key sensorimotor areas.

### 4.3 Inter-hemispheric connectivity differs as a function of subsequent motor recovery

Following intra-hemispheric connections, we next sought to evaluate the relevance of inter-hemispheric connections for post-stroke initial motor function and subsequent motor recovery. At the level of brain function, we observed recovery-related, but not function-related differences in inter-hemispheric disconnectivity. Specifically, we found that commissural motor connections were more disconnected in patients who didn’t recover, compared to patients who recovered, which is in line with a previous study (Yu et al., 2019). The importance of commissural connections for post-stroke motor recovery is corroborated by studies investigating the microarchitecture of the corpus callosum in the chronic phase post-stroke. These studies found that injury to the corpus callosum caused by stroke lesions through axonal degeneration correlated with poor subsequent motor recovery (J. Chen & Schlaug, 2013; Li et al., 2015; Radlinska et al., 2012; Stewart et al., 2017; Wang et al., 2012).

At the level of brain function, we found that inter-hemispheric connectivity was directed from the contralateral [ipsilesional] to the ipsilateral [contralesional] hemisphere in patients who recovered, while patients who didn’t recover showed the opposite pattern. The pattern observed in patients who didn’t recover, i.e., ipsilateral [contralesional] activity driving contralateral [ipsilesional] activity, can be interpreted in the light of the inter-hemispheric imbalance, related to decreased contralateral [ipsilesional] excitability and increased ipsilateral [contralesional] excitability (Nowak et al., 2009). As previous studies have focused almost exclusively on inter-hemispheric connectivity in M1 (Murase et al., 2004), the connectivity between the sensory areas is largely understudied ((Calautti et al., 2007), but see (Frías et al., 2018)). Combining tactile stimulation with high-quality MEG data, allowed us to investigate S1 and S2 in addition to M1. Our results suggest that the inter-hemispheric imbalance is not unique to M1 but extends to sensory areas. The inter-hemispheric imbalance framework informs non-invasive brain stimulation aiming at increasing contralateral [ipsilesional] excitability and decreasing ipsilateral [contralesional] excitability to enhance motor recovery post-stroke (Du et al., 2019; Lefebvre et al., 2013; Lindenberg et al., 2010; O’Shea et al., 2014; Ward & Cohen, 2004).

In summary, we found that differences in inter-hemispheric structural and functional connectivity relate to recovery-related rather than function-related processes. We found that patients who recovered had more intact commissural motor connections and functional connectivity directed from the contralateral [ipsilesional] to the ipsilateral [contralesional] hemisphere in acute brain imaging.

### 4.4 Predicting subsequent motor recovery post-stroke

Capitalising on the group differences between patients who recovered and patients who didn’t recover, we asked whether we could predict which of the low function patients will go on to achieve some recovery, based on very early post-stroke data. Generally, subsequent motor recovery is highly correlated to the initial function/impairment, however, this is not true for patients with initially severe upper limb impairment (Prabhakaran et al., 2008; Winters et al., 2015). Of these patients approximately half experience recovery while the other half do not (Winters et al., 2015). Therefore, there is a real need to improve outcome prediction in low function/more impaired patients. Here, we used properties from brain function and structure early post-stroke to predict whether low function patients were likely to recover or not. Model selection measures suggest a better model fit for the winning model, combining functional and structural features, compared to unimodal models. We found that, independent of initial severity, patients who had stronger functional inter-hemispheric connectivity and more intact motor projection connections were more likely to show subsequent motor recovery. Note that this prediction analysis focussed only on the subsample of initially low function patients. While this decision is mechanistically informed, it impacts the sample size. Further research focussing on the subsample of low function/more impaired patients are needed to fully uncover the mechanisms of recovery in this subsample.

### 4.5 Limitations and future directions

Post-stroke initial motor function/impairment and subsequent motor recovery, as well as the accompanying changes in brain function and structure, are multifarious and highly complex. While we focus on an important subset of brain functional and brain structural features, it is nevertheless a subset of an enormous feature space. Dimensionality reduction through feature selection is a common approach in outcome prediction models but usually relies on a priori decision making. Unsupervised high-dimensional methods (such as (Schrouff et al., 2013)) can overcome this problem but need extremely large datasets. As structural brain data are acquired as part of the clinical routine in some countries, large datasets of structural data post-stroke exist (such as (Liew et al., 2020)). However, functional brain data is not acquired routinely, and so there are no large datasets of functional brain data post stroke and large datasets combining brain structure and function.

As common practice, we collapsed right and left hemispheric strokes, however, the possibility of systematic differences in functional connectivity between left and right hemispheric strokes has been highlighted recently (Song et al., 2023). Collapsing across lesion side, as well as gender is, given the sample size, a pragmatic decision.

The functional and structural data used here were not acquired as part of clinical routines. However, to validate the results on large datasets and embed prediction models based on neuroimaging in clinical practice and care pathways the functional and structural features need to be extracted from inexpensive and accessible technology. To this end, high-quality MRI and MEG need to be replaced by routine scans and EEG. Strong correlation between MEG and EEG (Illman et al., 2020), as well as recent developments in mobile EEG (Niso et al., 2023) pave the way for low-cost assessment of functional measures at the bedside. Bedside recordings further allow the inclusion of underserved populations, i.e., patients with severe disabilities. Finally, to further improve prediction additional features, such as sensory impairment, other measurement depicting the quality of upper limb movement (Kwakkel et al., 2019), presence and strength of transcranial magnetic stimulation induced elicit motor-evoked potentials (Stinear et al., 2012), treatment type (i.e., thrombolysis or thrombectomy) stroke type (Grima et al., 2024), and several time points (acute and sub-acute stage) should be considered in future.

## Acknowledgments

We thank Suvi Heikkilä, Jari Kainulainen, Jyrki Mäkelä and Mia Illman for their help with MEG data acquisition. The authors thank the HUS occupational therapist for performing clinical testing.

## Author contributions

CZ: Conceptualization, Formal analysis, Methodology, Visualisation, Writing – original draft

NSW: Conceptualization, Funding acquisition, Writing – review and editing

NF: Conceptualization, Funding acquisition, Writing – review and editing

SV: Conceptualization, Writing – review and editing

AJQ: Methodology, Software, Writing – review and editing

EK: Data Curation, Project administration, Writing – original draft

KL: Conceptualization, Data Curation, Project administration, Writing – original draft

