## Supplemental Material for "Post-stroke changes in brain structure and function can both influence acute upper limb function and subsequent recovery"

**Supplemental Information – Table 1:** Clinical details of the patients.

| Study | Patient | Gender | Age | AH | Site | NHPT | |
|---|---|---|---|---|---|---|---|
| | | | | | | $T_0$ | $T_1$ |
| Laaksonen et al., 2012 | 1 | M | 60 | R | C | 120 | 101 |
| | 2 | F | 67 | L | S | 38 | 30 |
| | 3 | F | 72 | L | C | 33 | 24 |
| | 4 | F | 55 | R | CS | 120 | 120 |
| | 5 | M | 68 | L | CS | 58 | 34 |
| | 6 | F | 84 | R | C | 120 | 64 |
| | 7 | F | 68 | L | S | 120 | 120 |
| | 8 | F | 72 | R | CS | 120 | 120 |
| | 9 | M | 62 | L | CS | 47 | 43 |
| | 10 | F | 74 | R | S | 120 | 44 |
| | 11 | M | 78 | L | S | 120 | 30 |
| Parkkonen et al., 2018 | 1 | F | 59 | R | S | 180 | 180 |
| | 2 | M | 59 | R | C | 77 | 28 |
| | 3 | M | 57 | L | C | 180 | 55 |
| | 4 | M | 68 | R | S | 28 | 28 |
| | 5 | M | 71 | R | CS | 35 | 26 |
| | 6 | F | 59 | L | C | 35 | 22 |
| | 7 | M | 76 | L | CS | 29 | 23 |
| | 8 | M | 74 | L | S | 39 | 31 |
| | 9 | M | 66 | R | CS | 41 | 28 |
| | 10 | F | 68 | R | C | 32 | 21 |
| | 11 | F | 59 | R | S | 37 | 26 |
| | 12 | M | 45 | R | CS | 21 | 20 |
| | 13 | F | 58 | R | CS | 23 | 23 |
| | 14 | F | 66 | L | C | 180 | 180 |
| | 15 | F | 73 | L | CS | 180 | 131 |
| | 16 | F | 67 | R | CS | 42 | 28 |
| | 17 | M | 75 | R | CS | 180 | 180 |
| | 18 | F | 75 | L | S | 180 | 180 |
| | 19 | M | 64 | L | S | 180 | 180 |
| | 20 | M | 65 | L | S | 180 | 32 |
| | 21 | F | 74 | R | S | 180 | 180 |
| | 22 | M | 67 | R | CS | 180 | 180 |
| | 23 | M | 47 | R | CS | 180 | 180 |
| | 24 | F | 78 | R | CS | 180 | 180 |
| | 25 | M | 66 | R | S | 155 | 20 |

AH, affected hemisphere; R, Right; L, Left; C, cortical; CS, cortico-subcortical; S, subcortical; T0, 1–7 days; T1, 1 month

### Supplemental Information - Figures

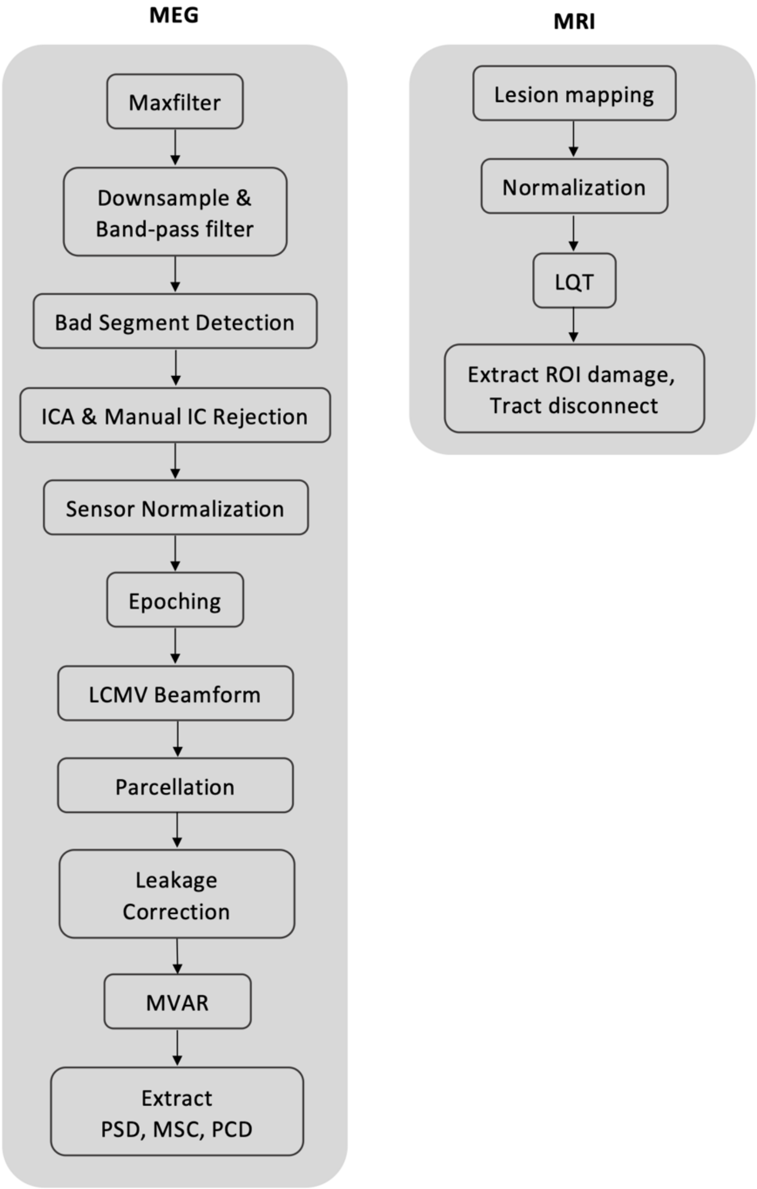


***SI Fig. 1.*** *A schematic for the processing pipeline.*

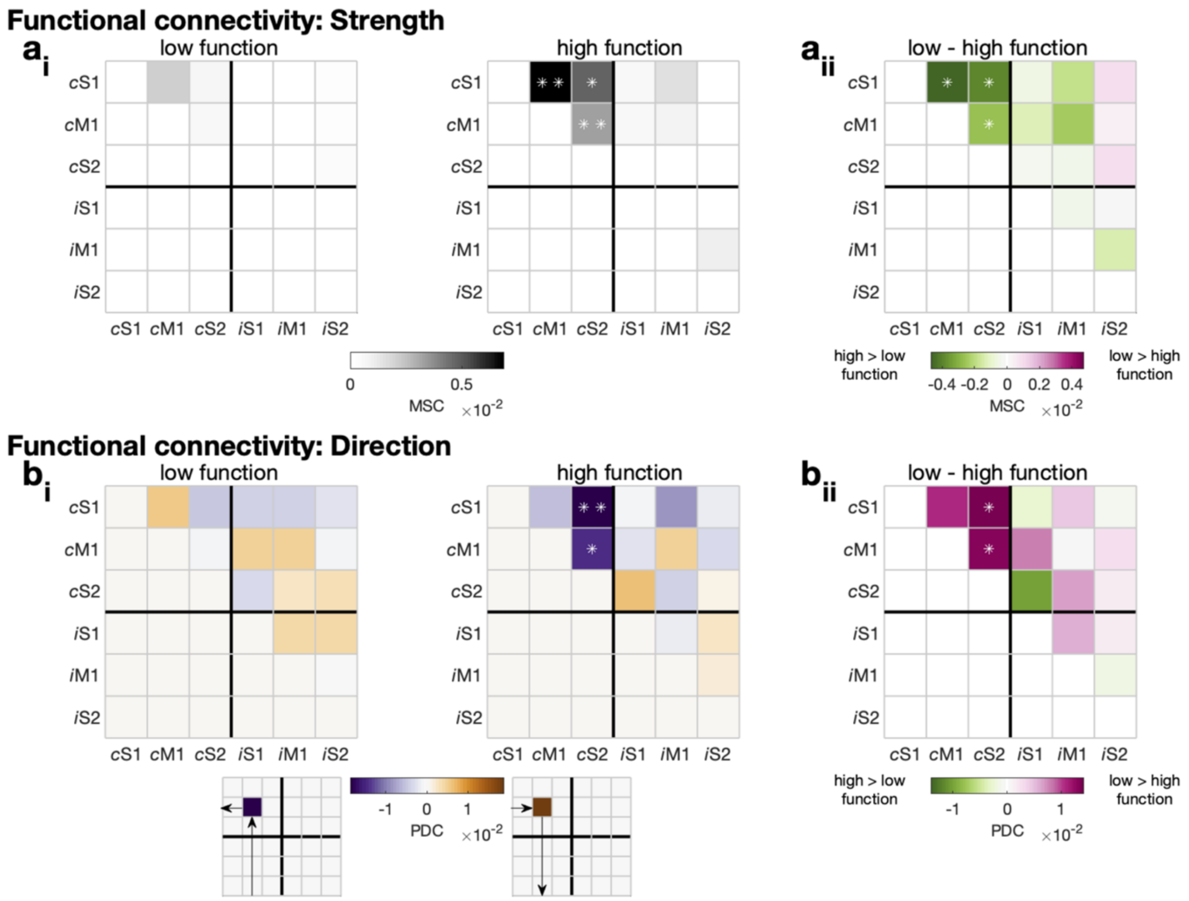


***SI Fig. 2.*** *Connectivity strength and direction for low function (left) and high function (right) patients.*
*ai) Connectivity strength for low function (left) and high function (right) patients. Connections, where the strength differs significantly from zero are highlighted (p<0.05 *, p<0.01**).*
*aii) Difference in connectivity strength between low and high function patients. Significant cells are highlighted (p<0.05 *, p<0.01**).*
*bi) Connectivity direction for low function (left) and high function (right) patients. The direction for purple cells is row to column, whereby the direction for brown cells is column to row (see also the legend below). Connections, where the direction differs significantly from zero, are highlighted (p<0.05 *, p<0.01**).*
*bii) Difference in connectivity direction between low and high function patients. Significant cells are highlighted (p<0.05 *, p<0.01**).*

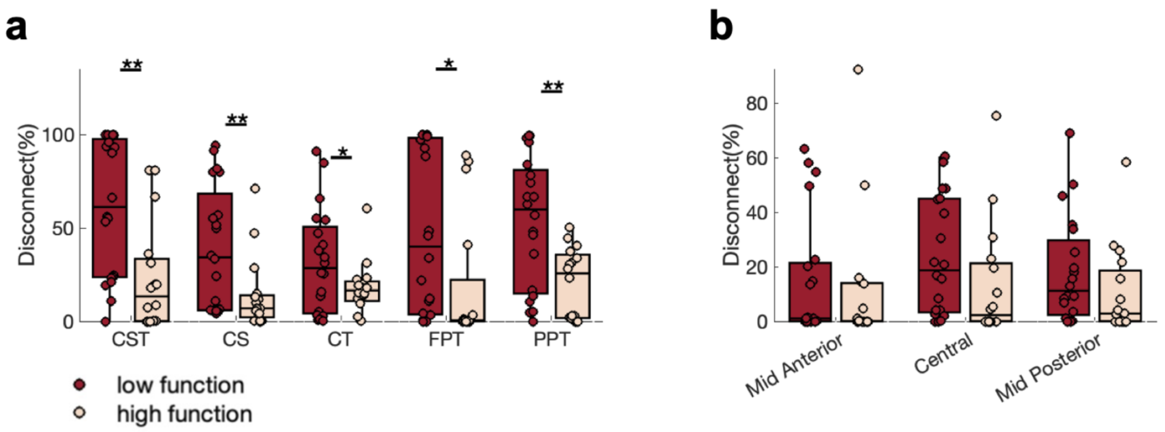


***SI Fig. 3.*** *Tract disconnect for low and high function patients.*
*a) Motor projection connections (corticospinal tract [CST], corticostriatal pathway [CS], corticothalamic pathway [CT], frontopontine tract [FPT], parietopontine tract [PPT]). Significant differences between groups are highlighted (p<0.05 *, p<0.01**).*
*b) Motor commissural connections (mid-anterior corpus callosum, central corpus callosum, mid-posterior corpus callosum). Significant differences between groups are highlighted (p<0.05 *, p<0.01**).*

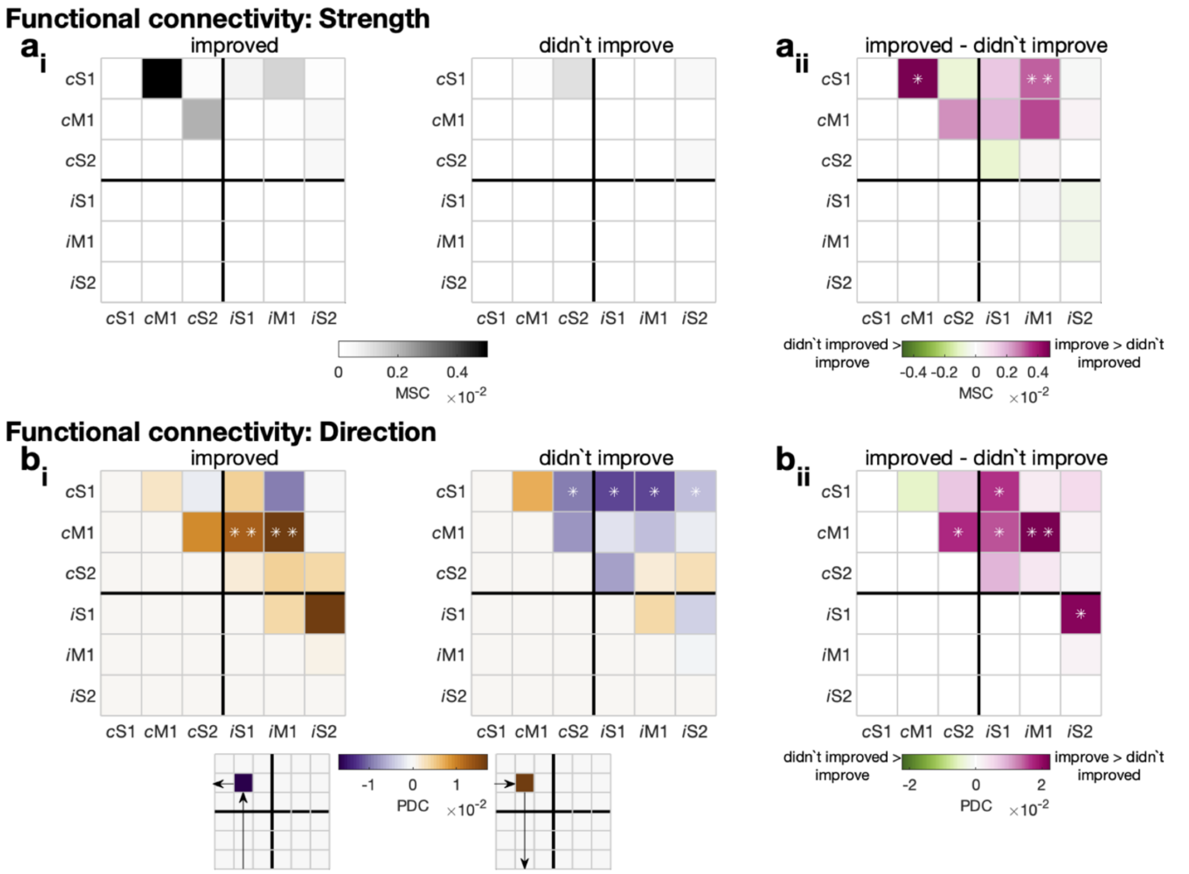


***SI Fig. 4.*** *Connectivity strength and direction patients who improved (left) and patients who didn't improve (right).*
*ai) Connectivity strength for patients who improved (left) and patients who didn't improve (right). Connections, where the strength differs significantly from zero are highlighted (p<0.05 *, p<0.01**).*
*aii) Difference in connectivity strength between patients who improved and patients who didn't improve. Significant cells are highlighted (p<0.05 *, p<0.01**).*
*bi) Connectivity direction for patients who improved (left) and patients who didn't improve (right) patients. The direction for purple cells is row to column, whereby the direction for brown cells is column to row (see also the legend below). Connections, where the direction differs significantly from zero, are highlighted (p<0.05 *, p<0.01**).*
*bii) Difference in connectivity direction between patients who improved and patients who didn't improve. Significant cells are highlighted (p<0.05 *, p<0.01**).*

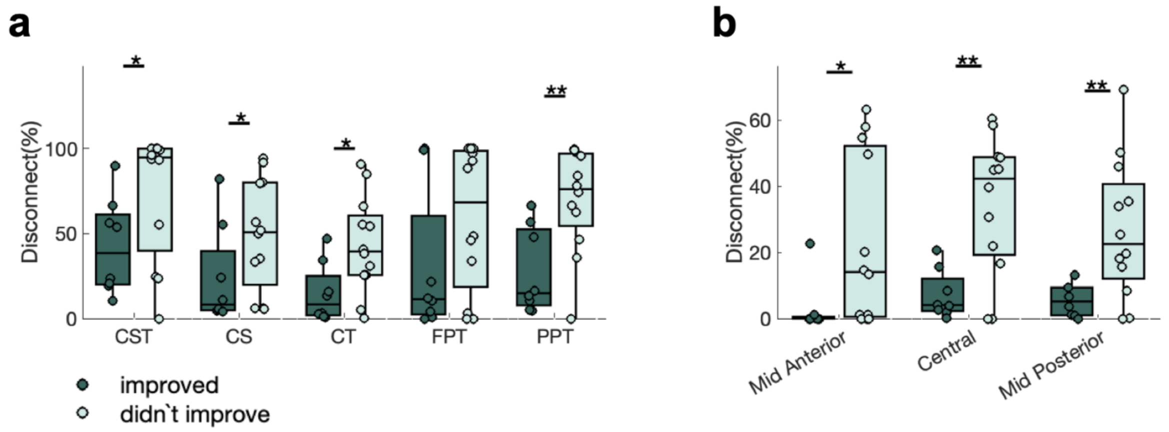


***SI Fig. 5.*** *Tract disconnect metrics for patients who improved and patients who didn't improve.*
*a) Motor projection connections (corticospinal tract [CST], corticostriatal pathway [CS], corticothalamic pathway [CT], frontopontine tract [FPT], parietopontine tract [PPT]). Significant differences between groups are highlighted (p<0.05 *, p<0.01**).*
*b) Motor commissural connections (mid-anterior corpus callosum, central corpus callosum, mid-posterior corpus callosum). Significant differences between groups are highlighted (p<0.05 *, p<0.01**).*

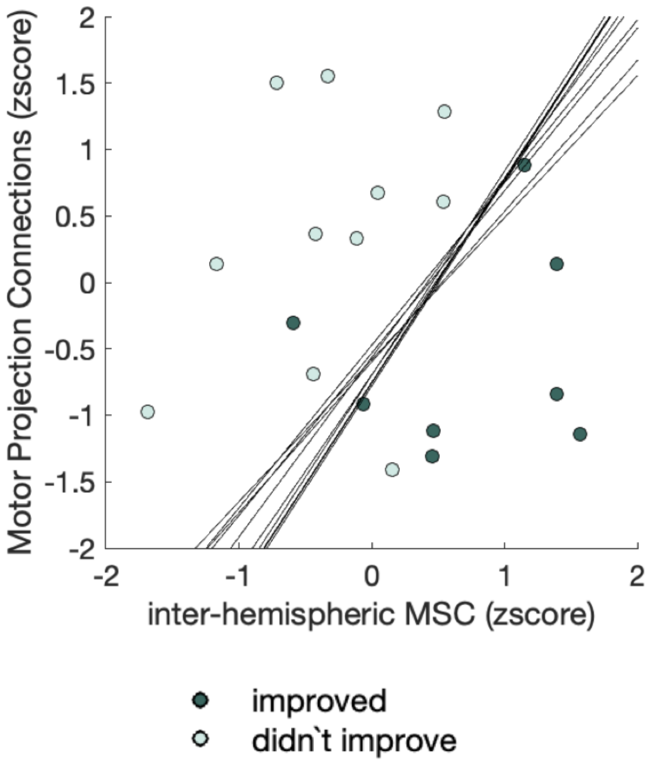


***SI Fig. 6.*** *Predictors included in the final logistic regression model. Each line represents the best separating hyperplane for the classifier for each fold of the leave one out cross validation.*

**Supplemental Information – Results**

Recovery-related differences in functional brain connectivity direction (Partial directed coherence, PDC)

| Group | From | To | df | t | p |
|---|---|---|---|---|---|
| improved | contralateral [ipsilesional] M1 | ipsilateral [contralesional] M1 | 7 | 3.97 | <0.01 |
| | contralateral [ipsilesional] M1 | ipsilateral [contralesional] S1 | 7 | 3.33 | <0.01 |
| didn't improve | contralateral [ipsilesional] S2 | contralateral [ipsilesional] S1 | 11 | -1.95 | <0.05 |
| | ipsilateral [contralesional] M1 | contralateral [ipsilesional] S1 | 11 | -1.93 | <0.05 |
| | ipsilateral [contralesional] S1 | contralateral [ipsilesional] S1 | 11 | -2.50 | <0.05 |
| | ipsilateral [contralesional] S2 | contralateral [ipsilesional] S1 | 11 | -2.62 | <0.01 |
| improved > didn't' improve | contralateral [ipsilesional] M1 | contralateral [ipsilesional] S2 | 18 | 1.97 | <0.05 |
| | contralateral [ipsilesional] M1 | ipsilateral [contralesional] M1 | 18 | 3.13 | <0.01 |
| | contralateral [ipsilesional] M1 | ipsilateral [contralesional] S1 | 18 | 2.10 | <0.05 |
| | contralateral [ipsilesional] S1 | ipsilateral [contralesional] S1 | 18 | 2.58 | <0.05 |
| | ipsilateral [contralesional] S1 | ipsilateral [contralesional] S2 | 18 | 2.08 | <0.05 |

**Supplemental Information – Results**

Model fit measures for the winning and two alternative models.

Inter-hemispheric MSC & Motor Projection Connections

Model Fit Measures

| | | | | | | | Overall Model Test | | |
|---|---|---|---|---|---|---|---|---|---|
| Model | Deviance | AIC | BIC | $R^2_{McF}$ | $R^2_{CS}$ | $R^2_N$ | $\chi^2$ | df | p |
| 1 | 13.3 | 19.3 | 22.2 | 0.508 | 0.495 | 0.669 | 13.7 | 2 | 0.001 |

CST

Model Fit Measures

| | | | | | | | Overall Model Test | | |
|---|---|---|---|---|---|---|---|---|---|
| Model | Deviance | AIC | BIC | $R^2_{McF}$ | $R^2_{CS}$ | $R^2_N$ | $\chi^2$ | df | p |
| 1 | 23.1 | 27.1 | 29.1 | 0.141 | 0.173 | 0.234 | 3.81 | 1 | 0.051 |

M1 and S1 β rebound

Model Fit Measures

| | | | | | | | Overall Model Test | | |
|---|---|---|---|---|---|---|---|---|---|
| Model | Deviance | AIC | BIC | $R^2_{McF}$ | $R^2_{CS}$ | $R^2_N$ | $\chi^2$ | df | p |
| 1 | 18.5 | 22.5 | 24.5 | 0.312 | 0.343 | 0.463 | 8.39 | 1 | 0.004 |

AIC Akaike information criterion
BIC Bayesian information criterion
$R^2_{MCF}$ McFadden's $R^2$
$R^2_{CS}$ Cox & Snell's $R^2$
$R^2_N$ Nagelkerke's $R^2$
$X^2$ Chi-square
